# Language impairment in motor neuron disease phenotypes different from classical amyotrophic lateral sclerosis: a review

**DOI:** 10.1101/2021.01.31.21250860

**Authors:** Benedetta Sbrollini, Alice Naomi Preti, Stefano Zago, Costanza Papagno, Ildebrando Marco Appollonio, Edoardo Nicolò Aiello

## Abstract

**Background:** Up to 35-40% of patients with amyotrophic lateral sclerosis (ALS) present with language deficits falling within the spectrum of frontotemporal degeneration (FTD). It is currently debated whether frontotemporal involvement occurs or not in motor neuron disease (MND) phenotypes that differ from classical ALS (*i*.*e*., both non-ALS MNDs and non-classical ALS endo-phenotypes) - this stance being supported by the notion of a common pathology underlying MNDs. To investigate whether language dysfunctions also occur in patients with different-from-classical-ALS MNDs can; a) help determine whether the MND-FTD *continuum* could be broadened at a neuropsychological level; b) convey relevant entailments to cognitive diagnostics in these populations.

**Aims:** The present study thus aimed at reviewing evidence regarding language impairment in different-from-classical-ALS MND patients. Preferred Reporting Items for Systematic Reviews and Meta-Analyses guidelines were consulted to implement and report the present review. Studies were included if a) language was quantitatively assessed b) in patients diagnosed with different-from-classical-ALS MND phenotypes. Studies assessing demented patients only were excluded. From an original *N*=1117 contributions, *N*=20 group studies were finally included. Secondary outcomes were taken into account for qualitatively assessing potential biases in generalizing results.

**Main contribution:** Studies were divided into those assessing predominant-upper vs. - lower MND patients (UMND/LMND). Language dysfunctions appeared to be more prevalent and severe in UMND patients. Language screeners were able to detect language deficits in both groups. Lexical-semantic deficits appeared to be highly prevalent in both groups and a selective difficulty in action-vs. object-naming was systematically detected. Morpho-syntactic deficits were seldom reported in both groups. Phonological deficits and central dysgraphic features were found in UMND patients only.

**Conclusion:** Patients with different-from-classical-ALS MND phenotypes display language deficits similar to those of classical ALS patients (as far as both prevalence and type are concerned) and thus could be validly included in the MND-FTD *continuum* at a neuropsychological level. A greater cortical involvement might account for language deficits being more severe in UMND patients. Consistently with guidelines for cognitive assessment in ALS patients, action-naming tasks might represent a valid and sensitive tool for assessing language in UMND/LMND patients too.

## 1. Introduction

Motor neuron diseases (MND) are phenotypically heterogeneous neurodegenerative syndromes affecting both motor and extra-motor systems [Al-Chalabi *et al*., 2016]. Frontotemporal involvement occurs due to genetic and pathophysiological links shared between frontotemporal degenerations (FTD) and MNDs [Zago *et al*., 2011; Burrell *et al*., 2016].

Phenotypic distinctions in MNDs mostly rely on motor descriptors – *i*.*e*., predominant-upper vs. -lower motor neuron involvement as well as spinal vs. bulbar affected sites [Eisen & Shaw, 2007]. Several others both qualitative and quantitative descriptors are also acknowledged (*e*.*g*., genetic susceptibility and progression rate, respectively), although they lead to less systematic classifications [Al-Chalabi *et al*., 2016]. Nevertheless, whether to regard MND phenotypes as discrete nosological entities is still debated [Turner, 2019].

Neuropsychological (NPs) deficits within the FTD *spectrum* affect up to 50% of patients with amyotrophic lateral sclerosis (ALS) [Strong *et al*., 2017] – the most prevalent MND phenotype [Logroscino *et al*., 2018]. However, little is known about frontotemporal involvement among both different ALS endo-phenotypes [Chiò *et al*., 2011] and different-from-classical-ALS MNDs [Eisen & Shaw, 2007]. If a common underlying pathology is assumed [Al-Chalabi *et al*., 2016; Burrell *et al*., 2016; Turner, 2019], FTD-mimicking NPs deficits can be reasonably expected in MND phenotypes that differ from classical ALS (*i*.*e*., both non-ALS MNDs and non-classical ALS endo-phenotypes) [De Vries *et al*., 2019a]. Multidisciplinary evidence would indeed hint at this last stance [Tartaglia *et al*., 2009; Geser *et al*., 2011; Kosaka *et al*., 2012; Prudlo *et al*., 2012; Cooper-Knock *et al*., 2014; De Vries *et al*., 2017; Gómez-Tortosa *et al*., 2017].

Up to 35-40% of ALS patients are estimated to display language deficits [Strong *et al*., 2017] within the *spectrum* of primary progressive aphasias (PPA) – encompassing phonological, lexical-semantic and morpho-syntactic components in both productive and receptive modalities, as well as in both oral and written ones [Pinto-Grau *et al*., 2018].

As being semiotically circumscribable and pathognomonic of FTD-*spectrum* cognitive disorders, language deficits can be accounted as a more sensitive and specific marker of a widespread cognitive decline in MND patients [Taylor *et al*., 2012; Tsermentseli *et al*., 2016].

Besides being theoretically relevant, to further profile cognitive functions in patients with different MND phenotypes also conveys clinical entailments. Indeed, cognitive impairment negatively influences MND patients’ both clinical and ecological management [Christidi *et al*., 2018; Huynh *et al*., 2020]. Therefore, it is crucial to identify markers of NPs decline in this population.

To investigate language deficits can thus represent a fecund *medium* in order to investigate the nature and prevalence of FTD-mimicking cognitive deficits in different MND phenotypes. Aim of the present study was thus to review evidence reporting language impairment in these populations.

## 2. Methods

### 2.1. Search strategy and study selection process

In order to implement this review, Preferred Reporting Items for Systematic Reviews and Meta-Analyses guidelines [Liberati *et al*. 2009] were consulted. Contributions published in English until June 2020 were searched in PubMed and Scopus databases by entering the following search terms: “primary lateral sclerosis” OR “progressive muscular atrophy” OR “flail arm syndrome” OR “flail leg syndrome” OR “progressive bulbar palsy” OR “pseudobulbar palsy” OR “upper motor neuron” OR “lower motor neuron” AND “neuropsych*” OR “cognit*” OR “language” OR “linguistic” OR “aphasia” [Pinto-Grau *et al*., 2018]. PubMed fields of search were the title and abstract; Scopus provided with an additional field of research - the keywords. In order for a contribution to be included, a) language had to be assessed through a quantitative examination and b) patients had to be clinically diagnosed with a MND phenotype different from classical “Charcot’s” ALS (either non-ALS MNDs or non-classical ALS variants) [Chiò *et al*., 2011].

Studies were excluded if they: a) did not explicitly investigate language functions; b) only assessed classical ALS patients; c) were reviews or meta-analyses; d) were case reports or case series; e) took into consideration demented patients only.

Study selection process is shown in Figure 1. One-thousand and one-hundred-seventeen contributions were screened within the title and the abstract; *N*=174 were selected for eligibility and thus read entirely. Three more articles possibly meeting inclusion criteria were identified through hand search; only one of them was included. Twenty group/cohort studies were thus eventually selected for inclusion.

**Figure 1.**
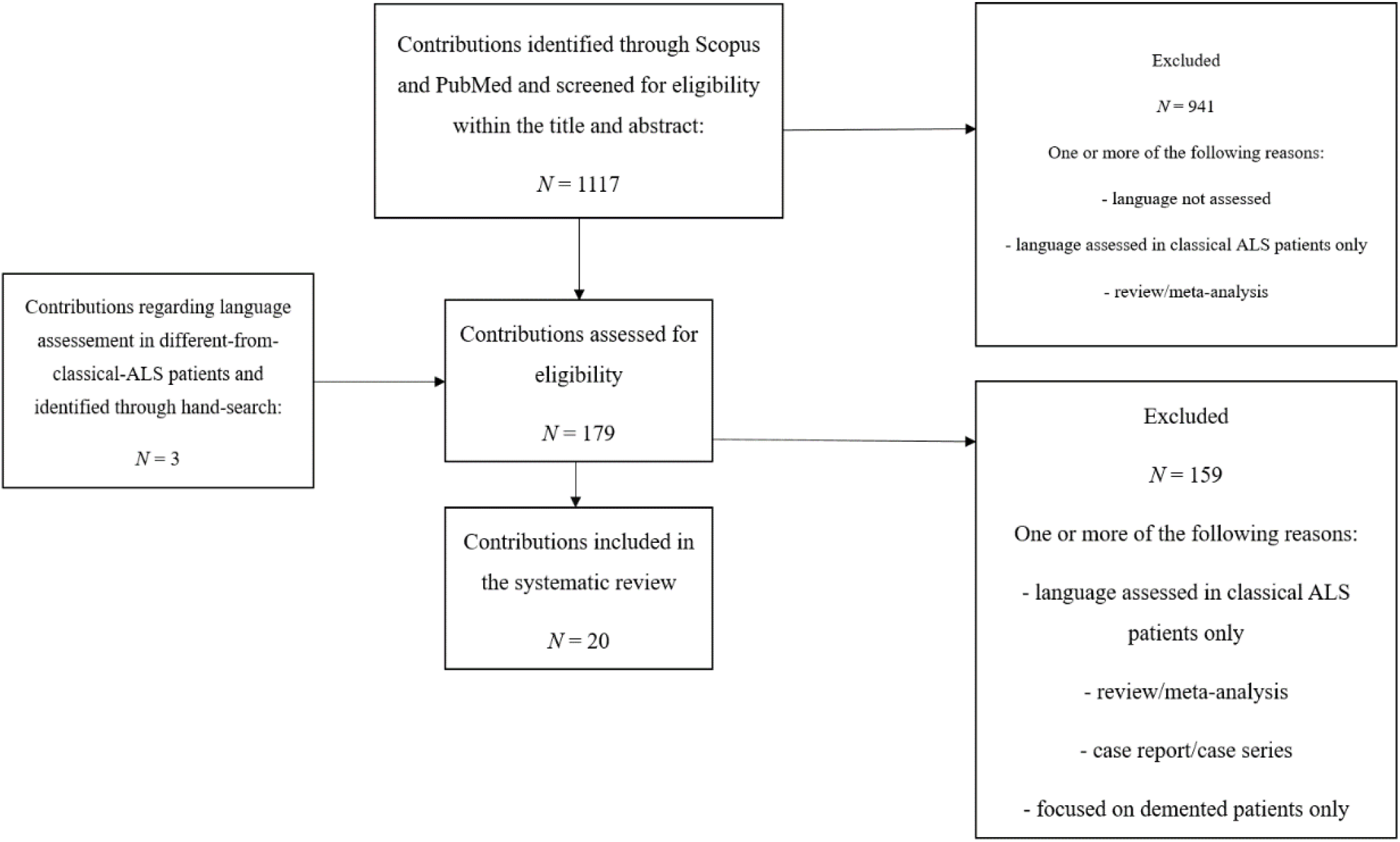
Diagram displaying study selection process (adapted from PRISMA guidelines). **Notes**. PRISMA=Preferred Reporting Items for Systematic Reviews and Meta-Analyses; MND=motor neuron disease.

### 2.2. Data extraction and considerations for bias management

The following language-related primary outcomes were extracted: language evaluation modalities; language components investigated; tasks/instruments adopted; comparison with healthy controls (HCs) and/or normative data; comparison with classical ALS patients; correlation between language and neural measures.

Both demographic and clinical outcomes were also taken into account: age, education, sex and language; presence of FTD; motor vs. neuropsychological onset [Mioshi *et al*., 2014]; disease duration; disease severity (as assessed by the ALS Functional Rating Scale – Revised [Cedarbaum *et al*., 1999]); presence of dysarthria. Potential biases in drawing inferences were qualitatively controlled by: a) taking into account the occurrence of dysarthria - which can mask or amplify linguistic deficits [Cobble, 1998]; b) assessing potential outliers in disease duration and severity measures [Al-Chalabi *et al*.,2016]; c) avoiding taking into account incidental neuro-anatomical/-functional measures; d) not regarding as language measures verbal fluency tasks [Aita *et al*., 2019]; e) not taking into account linguistic sub-scores of cognitive screeners [*e*.*g*., Folstein *et al*., 1975; Nasreddine *et al*. 2005] if they were not provided separately; f) taking into consideration comparisons between classical ALS patients and other MND phenotypes when estimating the prevalence of language impairment in the latter group.

## 3. Results

### 3.1. Outcomes overview

Patients’ demographic and clinical features are summarized in Table 1; Table 2 displays both primary and secondary outcomes. Results were grouped according to a motor descriptor – *i*.*e*., predominant-/pure-upper (*N*=12*) vs*. -lower (*N*=12) motor neuron involvement (UMND; LMND). Studies that investigated language in both UMND and LMND patients were regarded as two distinct contributions in the aforementioned counts.

**Table 1.**
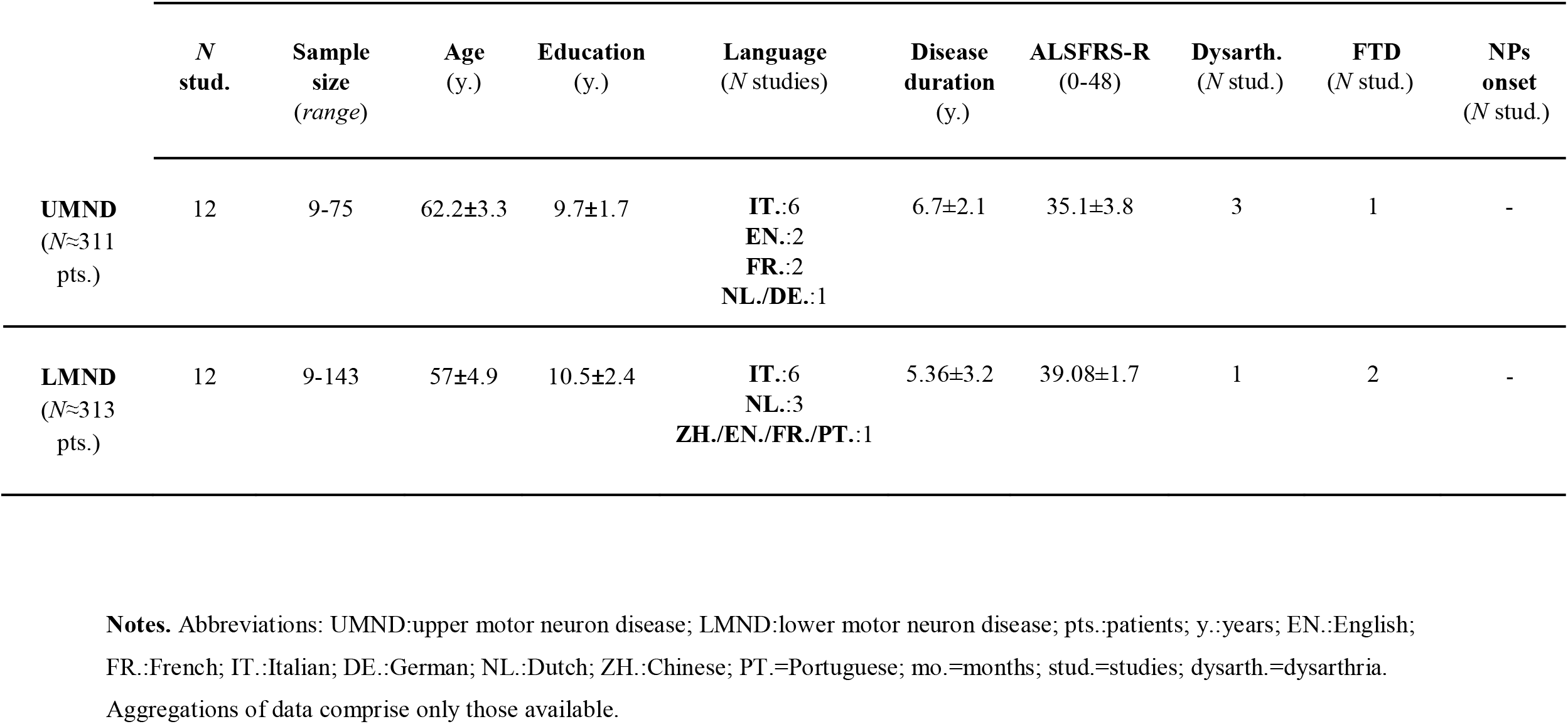
Synopsis of patients’ demographic and clinical features.

**Table 2.**
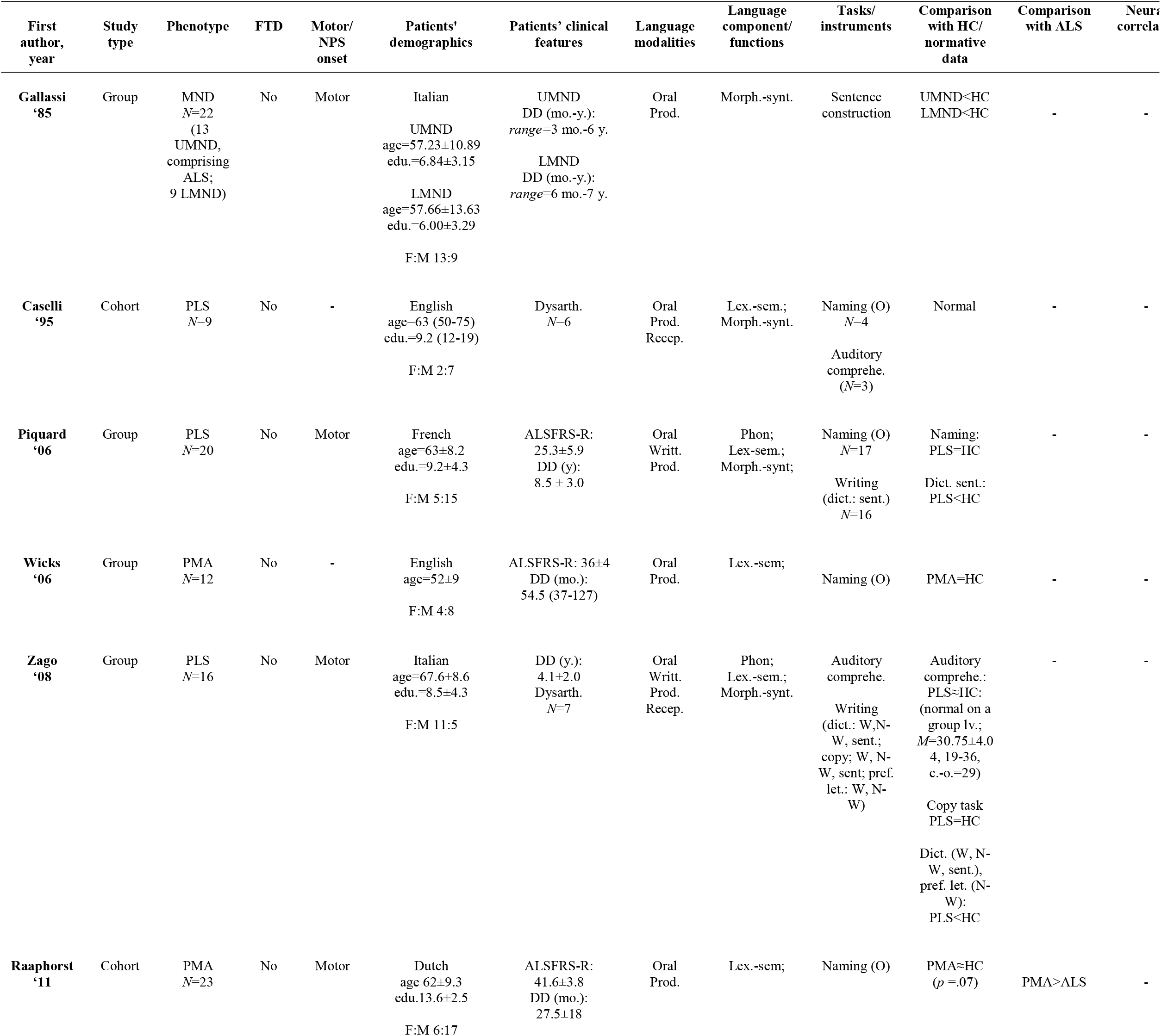
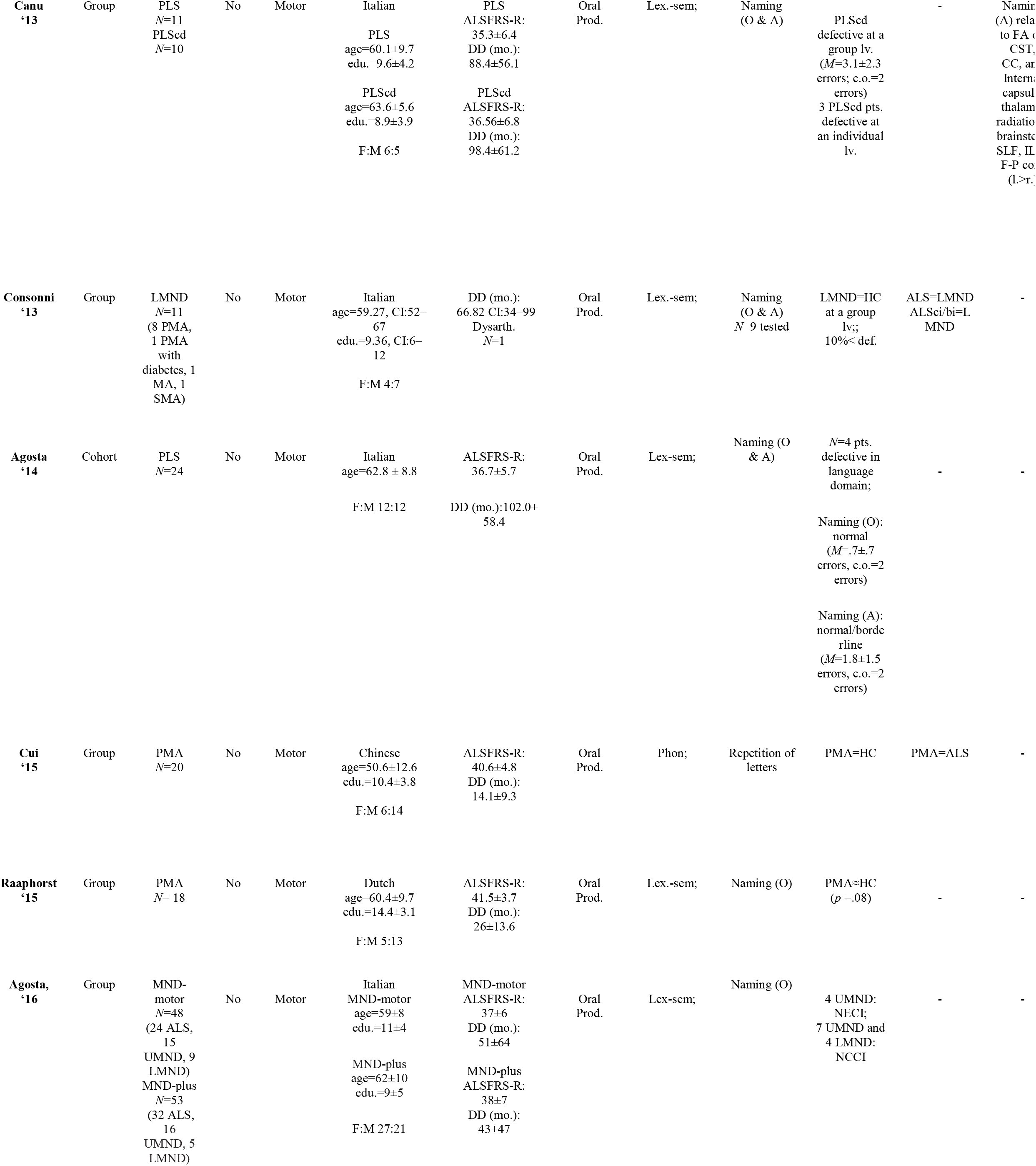
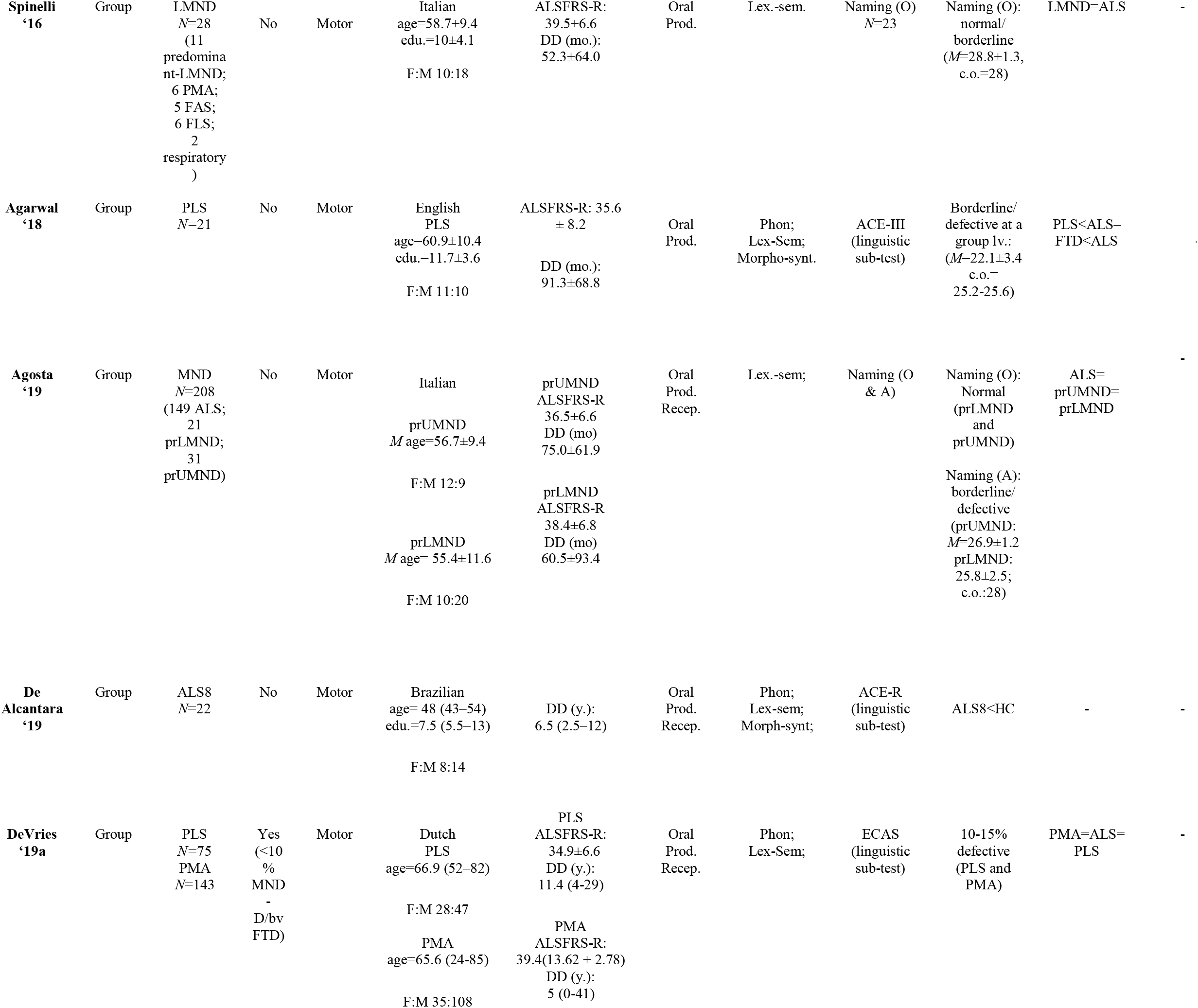
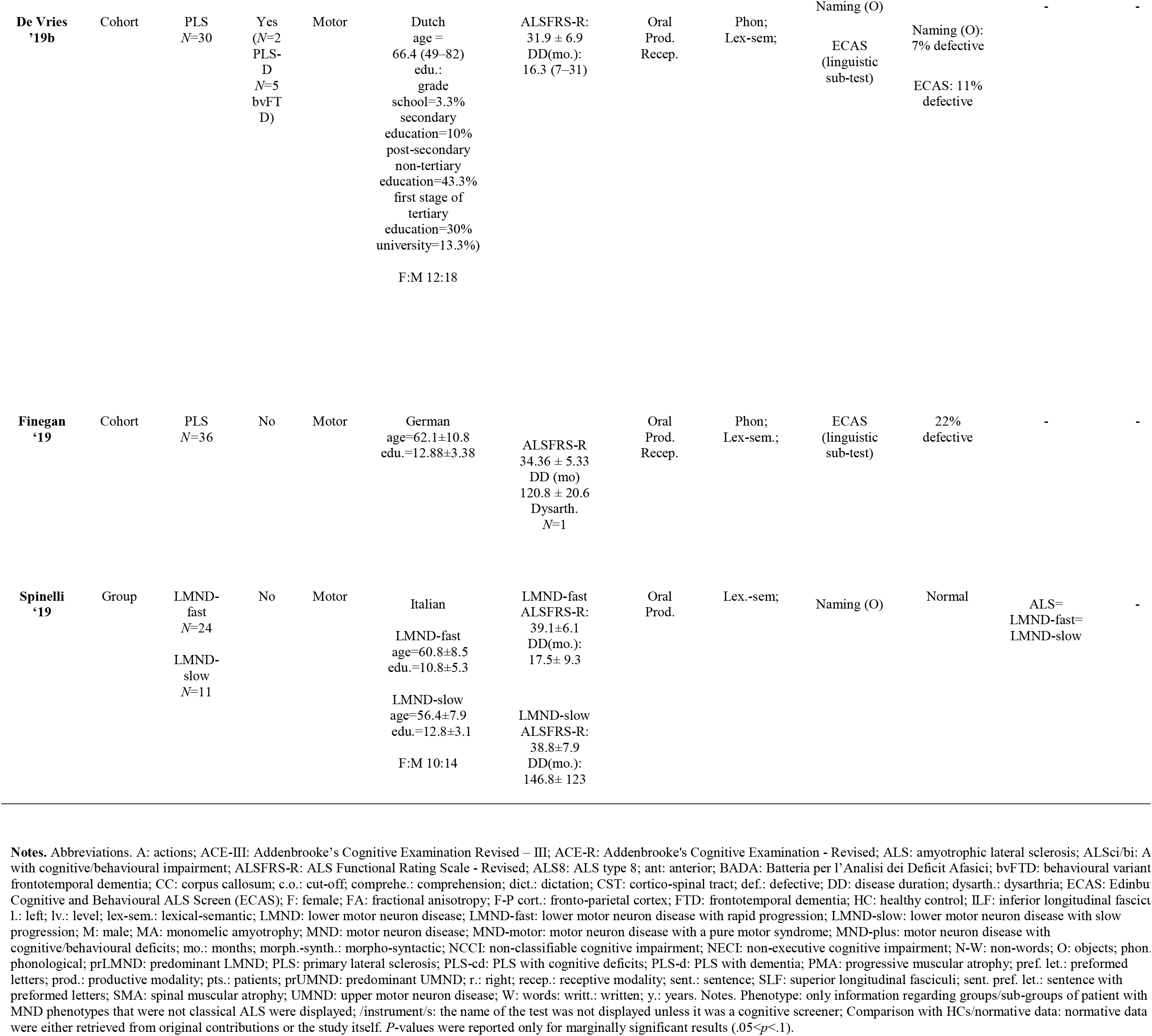
Summarization of extracted data.

| First author, year | Study type | Phenotype | FTD | Motor/ NPS onset | Patients' demographics | Patients' clinical features | Language modalities | Language component/ functions | Tasks/ instruments | Comparison with HC/ normative data | Comparison with ALS | Neurological correlations |
| --- | --- | --- | --- | --- | --- | --- | --- | --- | --- | --- | --- | --- |
| **Gallassi '85** | Group | MND<br>*N*=22<br>(13 UMND, comprising ALS; 9 LMND) | No | Motor | Italian<br>UMND<br>age=57.23±10.89<br>edu.=6.84±3.15<br>LMND<br>age=57.66±13.63<br>edu.=6.00±3.29<br>F:M 13:9 | UMND<br>DD (mo.-y.):<br>*range*=3 mo.-6 y.<br>LMND<br>DD (mo.-y.):<br>*range*=6 mo.-7 y. | Oral<br>Prod. | Morph.-synt. | Sentence construction | UMND<HC<br>LMND<HC | - | - |
| **Caselli '95** | Cohort | PLS<br>*N*=9 | No | - | English<br>age=63 (50-75)<br>edu.=9.2 (12-19)<br>F:M 2:7 | Dysarth.<br>*N*=6 | Oral<br>Prod.<br>Recep. | Lex.-sem.;<br>Morph.-synt. | Naming (O)<br>*N*=4<br>Auditory comprehe.<br>(*N*=3) | Normal | - | - |
| **Piquard '06** | Group | PLS<br>*N*=20 | No | Motor | French<br>age=63±8.2<br>edu.=9.2±4.3<br>F:M 5:15 | ALSFRS-R:<br>25.3±5.9<br>DD (y):<br>8.5 ± 3.0 | Oral<br>Writt.<br>Prod. | Phon;<br>Lex.-sem.;<br>Morph.-synt; | Naming (O)<br>*N*=17<br>Writing<br>(dict.: sent.)<br>*N*=16 | Naming:<br>PLS=HC<br>Dict. sent.:<br>PLS<HC | - | - |
| **Wicks '06** | Group | PMA<br>*N*=12 | No | - | English<br>age=52±9<br>F:M 4:8 | ALSFRS-R: 36±4<br>DD (mo.):<br>54.5 (37-127) | Oral<br>Prod. | Lex.-sem; | Naming (O) | PMA=HC | - | - |
| **Zago '08** | Group | PLS<br>*N*=16 | No | Motor | Italian<br>age=67.6±8.6<br>edu.=8.5±4.3<br>F:M 11:5 | DD (y.):<br>4.1±2.0<br>Dysarth.<br>*N*=7 | Oral<br>Writt.<br>Prod.<br>Recep. | Phon;<br>Lex.-sem.;<br>Morph.-synt. | Auditory comprehe.<br>Writing<br>(dict.: W,N-W, sent.;<br>copy; W, N-W, sent; pref.<br>let.: W, N-W) | Auditory comprehe.:<br>PLS≈HC:<br>(normal on a group 1v.;<br>*M*=30.75±4.0<br>4, 19-36,<br>c.-o.=29)<br>Copy task<br>PLS=HC<br>Dict. (W, N-W, sent.),<br>pref. let. (N-W):<br>PLS<HC | - | - |
| **Raaphorst '11** | Cohort | PMA<br>*N*=23 | No | Motor | Dutch<br>age 62±9.3<br>edu.13.6±2.5<br>F:M 6:17 | ALSFRS-R:<br>41.6±3.8<br>DD (mo.):<br>27.5±18 | Oral<br>Prod. | Lex.-sem; | Naming (O) | PMA≈HC<br>(*p* =.07) | PMA>ALS | - |
| Canu<br>'13 | Group | PLS<br>*N*=11<br>PLScd<br>*N*=10 | No | Motor | Italian<br>PLS<br>age=60.1±9.7<br>edu.=9.6±4.2<br>PLScd<br>age=63.6±5.6<br>edu.=8.9±3.9<br>F:M 6:5 | PLS<br>ALSFRS-R:<br>35.3±6.4<br>DD (mo.):<br>88.4±56.1<br>PLScd<br>ALSFRS-R:<br>36.56±6.8<br>DD (mo.):<br>98.4±61.2 | Oral<br>Prod. | Lex.-sem; | Naming<br>(O & A) | PLScd<br>defective at a<br>group lv.<br>(*M*=3.1±2.3<br>errors; c.o.=2<br>errors)<br>3 PLScd pts.<br>defective at<br>an individual<br>lv. | - | Namin<br>(A) rela<br>to FA c<br>CST,<br>CC, an<br>Intern;<br>capsul<br>thalam<br>radiatio<br>brainste<br>SLF, IL<br>F-P co<br>(1.>r. |
| Consonni<br>'13 | Group | LMND<br>*N*=11<br>(8 PMA,<br>1 PMA<br>with<br>diabetes, 1<br>MA, 1<br>SMA) | No | Motor | Italian<br>age=59.27, CI:52–<br>67<br>edu.=9.36, CI:6–<br>12<br>F:M 4:7 | DD (mo.):<br>66.82 CI:34–99<br>Dysarth.<br>*N*=1 | Oral<br>Prod. | Lex.-sem; | Naming<br>(O & A)<br>*N*=9 tested | LMND=HC<br>at a group<br>lv;;<br>10%< def. | ALS=LMND<br>ALSci/bi=L<br>MND | - |
| Agosta<br>'14 | Cohort | PLS<br>*N*=24 | No | Motor | Italian<br>age=62.8 ± 8.8<br>F:M 12:12 | ALSFRS-R:<br>36.7±5.7<br>DD (mo.):102.0±<br>58.4 | Oral<br>Prod. | Lex-sem; | Naming (O<br>& A) | *N*=4 pts.<br>defective in<br>language<br>domain;<br>Naming (O):<br>normal<br>(*M*=.7±.7<br>errors, c.o.=2<br>errors)<br>Naming (A):<br>normal/borde<br>rline<br>(*M*=1.8±1.5<br>errors, c.o.=2<br>errors) | - | - |
| Cui<br>'15 | Group | PMA<br>*N*=20 | No | Motor | Chinese<br>age=50.6±12.6<br>edu.=10.4±3.8<br>F:M 6:14 | ALSFRS-R:<br>40.6±4.8<br>DD (mo.):<br>14.1±9.3 | Oral<br>Prod. | Phon; | Repetition of<br>letters | PMA=HC | PMA=ALS | - |
| Raaphorst<br>'15 | Group | PMA<br>*N*= 18 | No | Motor | Dutch<br>age=60.4±9.7<br>edu.=14.4±3.1<br>F:M 5:13 | ALSFRS-R:<br>41.5±3.7<br>DD (mo.):<br>26±13.6 | Oral<br>Prod. | Lex.-sem; | Naming (O) | PMA≈HC<br>(*p* =.08) | - | - |
| Agosta,<br>'16 | Group | MND-motor<br>*N*=48<br>(24 ALS,<br>15<br>UMND, 9<br>LMND)<br>MND-plus<br>*N*=53<br>(32 ALS,<br>16<br>UMND, 5<br>LMND) | No | Motor | Italian<br>MND-motor<br>age=59±8<br>edu.=11±4<br>MND-plus<br>age=62±10<br>edu.=9±5<br>F:M 27:21 | MND-motor<br>ALSFRS-R:<br>37±6<br>DD (mo.):<br>51±64<br>MND-plus<br>ALSFRS-R:<br>38±7<br>DD (mo.):<br>43±47 | Oral<br>Prod. | Lex-sem; | Naming (O) | 4 UMND:<br>NECI;<br>7 UMND and<br>4 LMND:<br>NCCI | - | - |
| **Spinelli**<br>**'16** | Group | LMND<br>*N*=28<br>(11<br>predomina<br>nt-LMND;<br>6 PMA;<br>5 FAS;<br>6 FLS;<br>2<br>respiratory<br>) | No | Motor | Italian<br>age=58.7±9.4<br>edu.=10±4.1<br>F:M 10:18 | ALSFRS-R:<br>39.5±6.6<br>DD (mo.):<br>52.3±64.0 | Oral<br>Prod. | Lex.-sem. | Naming (O)<br>*N*=23 | Naming (O):<br>normal/<br>borderline<br>(*M*=28.8±1.3,<br>c.o.=28) | LMND=ALS | - |
| **Agarwal**<br>**'18** | Group | PLS<br>*N*=21 | No | Motor | English<br>PLS<br>age=60.9±10.4<br>edu.=11.7±3.6<br>F:M 11:10 | ALSFRS-R: 35.6<br>± 8.2<br>DD (mo.):<br>91.3±68.8 | Oral<br>Prod. | Phon;<br>Lex-Sem;<br>Morpho-synt. | ACE-III<br>(linguistic<br>sub-test) | Borderline/<br>defective at a<br>group lv.:<br>(*M*=22.1±3.4<br>c.o.=<br>25.2-25.6) | PLS<ALS–<br>FTD<ALS | . |
| **Agosta**<br>**'19** | Group | MND<br>*N*=208<br>(149 ALS;<br>21<br>prLMND;<br>31<br>prUMND) | No | Motor | Italian<br>prUMND<br>*M* age=56.7±9.4<br>F:M 12:9<br>prLMND<br>*M* age= 55.4±11.6<br>F:M 10:20 | prUMND<br>ALSFRS-R<br>36.5±6.6<br>DD (mo)<br>75.0±61.9<br>prLMND<br>ALSFRS-R<br>38.4±6.8<br>DD (mo)<br>60.5±93.4 | Oral<br>Prod.<br>Recep. | Lex.-sem; | Naming (O<br>& A) | Naming (O):<br>Normal<br>(prLMND<br>and<br>prUMND)<br>Naming (A):<br>borderline/<br>defective<br>(prUMND:<br>*M*=26.9±1.2<br>prLMND:<br>25.8±2.5;<br>c.o.:28) | ALS=<br>prUMND=<br>prLMND | - |
| **De**<br>**Alcantara**<br>**'19** | Group | ALS8<br>*N*=22 | No | Motor | Brazilian<br>age= 48 (43–54)<br>edu.=7.5 (5.5–13)<br>F:M 8:14 | DD (y.):<br>6.5 (2.5–12) | Oral<br>Prod.<br>Recep. | Phon;<br>Lex-sem;<br>Morph-synt; | ACE-R<br>(linguistic<br>sub-test) | ALS8<HC | - | - |
| **DeVries**<br>**'19a** | Group | PLS<br>*N*=75<br>PMA<br>*N*=143 | Yes<br>(<10<br>%<br>MND<br>-<br>D/bv<br>FTD) | Motor | Dutch<br>PLS<br>age=66.9 (52–82)<br>F:M 28:47<br>PMA<br>age=65.6 (24-85)<br>F:M 35:108 | PLS<br>ALSFRS-R:<br>34.9±6.6<br>DD (y.):<br>11.4 (4-29)<br>PMA<br>ALSFRS-R:<br>39.4(13.62 ± 2.78)<br>DD (y.):<br>5 (0-41) | Oral<br>Prod.<br>Recep. | Phon;<br>Lex-Sem; | ECAS<br>(linguistic<br>sub-test) | 10-15%<br>defective<br>(PLS and<br>PMA) | PMA=ALS=<br>PLS | - |
| **De Vries '19b** | Cohort | PLS<br>*N*=30 | Yes<br>(*N*=2<br>PLS-D<br>*N*=5<br>bvFTD) | Motor | Dutch<br>age =<br>66.4 (49–82)<br>edu.:<br>grade<br>school=3.3%<br>secondary<br>education=10%<br>post-secondary<br>non-tertiary<br>education=43.3%<br>first stage of<br>tertiary<br>education=30%<br>university=13.3%)<br>F:M 12:18 | ALSFRS-R:<br>31.9 ± 6.9<br>DD(mo.):<br>16.3 (7–31) | Oral<br>Prod.<br>Recep. | Phon;<br>Lex-sem; | Naming (O)<br>ECAS<br>(linguistic<br>sub-test) | Naming (O):<br>7% defective<br>ECAS: 11%<br>defective | - | - |
| **Finegan '19** | Cohort | PLS<br>*N*=36 | No | Motor | German<br>age=62.1±10.8<br>edu.=12.88±3.38 | ALSFRS-R<br>34.36 ± 5.33<br>DD (mo)<br>120.8 ± 20.6<br>Dysarth.<br>*N*=1 | Oral<br>Prod.<br>Recep. | Phon;<br>Lex-sem.; | ECAS<br>(linguistic<br>sub-test) | 22%<br>defective | - | - |
| **Spinelli '19** | Group | LMND-fast<br>*N*=24<br>LMND-slow<br>*N*=11 | No | Motor | Italian<br>LMND-fast<br>age=60.8±8.5<br>edu.=10.8±5.3<br>LMND-slow<br>age=56.4±7.9<br>edu.=12.8±3.1<br>F:M 10:14 | LMND-fast<br>ALSFRS-R:<br>39.1±6.1<br>DD(mo.):<br>17.5± 9.3<br>LMND-slow<br>ALSFRS-R:<br>38.8±7.9<br>DD(mo.):<br>146.8± 123 | Oral<br>Prod. | Lex.-sem; | Naming (O) | Normal | ALS=<br>LMND-fast=<br>LMND-slow | - |

Both receptive and productive modalities were assessed. All studies included oral language evaluation, whereas only two also investigated writing abilities. The most investigated component was the lexical-semantic (*N*=18) one, followed by phonological (*N*=8) and morpho-syntactic *N* =6) ones.

Frequently administered tests included the Boston Naming Test (BNT) [Kaplan *et al*., 1983], the oral confrontation object- and action-naming sub-tests from the Batteria per l’Analisi dei Deficit Afasici (BADA) [Miceli *et al*., 1991] and the Token Test (TT) [De Renzi & Faglioni, 1978]. Cognitive screeners including language sub-tests were the Addenbrooke’s Cognitive Examination – Revised/-III (ACE-R/-III) [Mioshi *et al*., 2006; Hsieh *et al*., 2013] and the Edinburgh Cognitive and Behavioral ALS Screen (ECAS) [Abrahams *et al*., 2014]. The ACE-R linguistic sub-test assesses both productive and receptive language, as well as reading and writing abilities. The ECAS linguistic sub-test encompasses confrontation naming, single-word comprehension and spelling tasks. Therefore, the ACE-R/-III linguistic sub-test was regarded as a measure of phonological, lexical-semantic and morpho-syntactic components, whereas the ECAS linguistic sub-test as a measure of phonological and lexical-semantic components.

### 3.2. Language functioning in UMND patients

#### 3.2.1. Phonological and morpho-syntactic components in UMND patients

Hints at phonology being possibly impaired in UMND patients come from the evaluation of their writing abilities. Piquard *et al*. (2006) found that primary lateral sclerosis (PLS) patients made spelling errors in a task of writing sentences to dictation [Goodglass & Kaplan, 1972]. Zago *et al*. (2008) also showed that PLS patients performed worse than HCs in writing to dictation of words, non-words and sentences, as well as in writing with preformed letters; however, PLS patients did not perform worse than HCs in a copy task (of words, non-words and sentences).

Investigations on morpho-syntactic abilities in patients with UMND phenotypes have yielded mixed results. Gallassi *et al*. (1985) found that MND patients with predominant UMN involvement (prUMND) performed worse on a sentence construction task when compared to both predominant-lower MND (prLMND) patients and HCs. It has nonetheless to be noted that ALS patients were included within the UMN group in this study.

Similarly, Piquard *et al*. (2006) reported that PLS patients showed more grammatical errors than HCs in the aforementioned sentence dictation task. Zago *et al*. (2008) also administered the TT to PLS patients, who proved to perform within the normal range as a group. Nonetheless, by taking into account the group mean combined with its standard deviation and the range, it can be inferred that some patients performed defectively at an individual level.

#### 3.2.2. Lexical-semantic components in UMND patients

Caselli *et al*. (1995) found that PLS patients performed normally on the BNT. Similarly, Piquard *et al*. (2006) reported that PLS patients performed comparably to HCs on a confrontation naming test [Deloche *et al*., 1997]. Nonetheless, Canu *et al*. (2013) found that, when compared to cognitively-unimpaired PLS patients, cognitively-impaired ones performed worse on the action-naming sub-test of the BADA, whereas not on its object-naming task. The Authors also found a significant left-greater-than-right association between damages to cortical/sub-cortical language-related white matter structures and action-naming scores in the PLS cohort. Similarly, Agosta *et al*. (2014) described a cohort of PLS patients who performed normally on the BADA object-naming sub-test, whilst at a borderline level on its action-naming task. The Authors also reported that *N*=4 PLS patients were classified as having language impairment based on the aforementioned measures.

In a subsequent work, the Authors [Agosta *et al*., 2016] aimed at comparing “pure” MND patients (*i*.*e*., who were classified as being cognitively-normal) to MND patients with neuropsychological deficits (MND-plus). Within each group, the Authors sub-divided patients according to motor phenotypes [Chiò *et al*., 2011] into classical ALS, pure LMND (PLMND) and pure UMND (PUMND) patients. Language was assessed through the object-naming task from the BADA. Authors also classified MND-plus patients according to their cognitive profiles: if dysexecutive symptoms were predominant (“executive cognitive impairment”, ECI); if dysfunctions of instrumental domains were predominant (“non-executive cognitive impairment”, NECI); if both executive and instrumental deficits were detectable (“non-classifiable cognitive impairment”, NCCI). Since *N*=4 UMND patients were classified as NECI, whereas *N*=7 as NCCI, it cannot be ruled out that some of them presented with language impairment. In a third study, the Authors [Agosta *et al*. 2019] administered BADA object- and action-naming sub-tests to a large cohort of MND patients including classical ALS, prUMND and prLMND patients [Chiò *et al*., 2011]. prUMND patients as a group proved to perform within the normal range on the object naming test, whereas it could be descriptively inferred that some of them performed defectively at an individual level. Nonetheless, when compared to both classical ALS and prLMND patients, prUMND patients did not prove to perform differently on either object- or action-naming tasks.

Inconsistently with Caselli *et al*.’s (1995) findings, De Vries *et al*. (2019b) found that 7% of their PLS cohort was defective on the BNT.

#### 3.2.3. Global language functioning in UMND patients

Agarwal *et al*. (2018) assessed language functions of PLS patients by means of the linguistic sub-test of the ACE-III [Hsieh *et al*., 2013]. By comparing the group-level performance (22.1±3.4) to normative data (95% CI [25.2, 25.6]) [The University of Sydney, n.d.] it can be inferred that patients performed defectively at least to an extent. Moreover, the Authors found that PLS patients as a group performed worse than ALS and ALS-FTD patients as well. De Vries *et al*. (2019b) found that 11% of their cohort of PLS patients performed abnormally on the ECAS linguistic-sub-test. A subsequent investigation by the same Authors [De Vries *et al*., 2019a] found that the prevalence for language deficits (as assessed by the ECAS linguistic sub-test) ranged between 10% and 15%. Finegan *et al*. (2019) reported that, among the cognitive domains assessed by the ECAS, the higher prevalence of deficits was found with regard to the linguistic sub-test (22% of the sample).

### 3.3. Language functioning in LMND patients

#### 3.3.1. Phonological and morpho-syntactic components in LMND patients

Morpho-syntactic abilities in LMND patients were specifically assessed by Gallassi *et al*. (1985) only, who reported that prLMND patients performed better in a phrase construction task when compared to both prUMND patients (which included classical ALS patients) and HCs. Similarly, one contribution only investigated phonological aspects [Cui *et al*., 2015]: the Authors compared a cohort of PMA patients to both classical ALS patients and HCs on a letter repetition task and did not detect significant differences between the three groups.

#### 3.3.2. Lexical-semantic components in LMND patients

Wicks *et al*. (2006) compared PMA patients to HCs on a naming task and did not find a significant between-group difference. Raaphorst *et al*. (2011) found a marginally-significant difference between PMA patients and HCs when the two groups were compared on the BNT – with patients performing slightly worse than HCs. In a subsequent contribution by the same Authors [Raaphorst *et al*., 2015], the previous results were confirmed – a marginally-significant trend towards a worse performance in PMA patients was found. Consonni *et al*. (2013) administered the BADA confrontation naming sub-tests (of objects and actions) to a phenotypically-heterogeneous cohort of patients affected by LMNDs. The Authors found that LMND patients were comparable to HCs on both tasks, although a small percentage of patients (less than 10%) showed abnormal naming scores – with no reported difference between objects and actions. Furthermore, on both tests, LMND patients performed comparably to both neuropsychologically-unimpaired and -impaired classical ALS patients. In the study by Agosta *et al*. (2016) [see above, 3.2.2.], *N*=4 PLMND patients were classified as NCCI – this suggesting that at least some of these patients might have presented with deficits in the language domain. Spinelli *et al*. (2016) found that LMND patients proved to perform within the normal range as a group on the object-naming task of the BADA – although the mean scores being just above the established cut-off (28) suggest that some of the patients might have presented with borderline/defective scores. Furthermore, a comparison between LMND and classical ALS patients on the object-naming task did not yield a significant difference. Similarly to the study by Consonni *et al*. (2013), Spinelli *et al*.’s (2016) investigation took into account highly heterogeneous patients as far as etiology is concerned. In the study by Agosta *et al*. (2019) [see above, 3.2.2.], prLMND patients proved to perform normally as a group on the object-naming task of the BADA, whereas defective individual-level performances could be inferred if comparing mean action-naming scores (25.8±2.5) to the original cut-off value – *i*.*e*., 28 [Miceli *et al*., 1991]. Furthermore, no significant differences between prLMND, prUMND and classical ALS patients on both naming tasks were detected. Spinelli *et al*. (2019) investigated language functions in LMND patients *via* the BADA object-naming. Patients were sub-divided into slowly and rapidly progressive. Both LMND groups proved to perform normally on the object-naming test, and their performance was comparable to a group of classical ALS patients.

#### 3.3.3. Global language functioning in UMND patients

De Alcântara *et al*. (2019) reported that patients affected with ALS8 – a familial form of MND characterized by a predominant involvement of LMNs – performed worse than HCs on the ACE-R linguistic sub-test. Moreover, De Vries *et al*. (2019b) found that a percentage ranging between 10% and 15% of PMA patients that underwent the ECAS showed abnormal scores on its linguistic sub-test. Furthermore, no between-group differences were found on the ECAS linguistic sub-test between PMA, PLS and ALS patients.

## 4. Discussion

### 4.1. Theoretical and clinical entailments

Overall, the present review suggests that language dysfunctions can occur in patients affected by MND phenotypes different from classical ALS, as well as that the nature of language deficits in patients diagnosed with different phenotypes of MND may be similar [*e*.*g*., De Vries *et al*., 2019a]. Indeed, the language profile of UMND/LMND patients appears to notably overlap with that of classical ALS patients [Pinto-Grau *et al*., 2018]. It can be thus reasonably speculated that also UMND/LMND patients could be validly included in the MND-FTD *continuum* at a neuropsychological level [De Vries *et al*., 2019a; Turner, 2019]. Consistently, frontotemporal systems involvement in different-from-classical-ALS patients [*e*.*g*., Saberi *et al*., 2015; Pinto *et al*., 2019] appears to also be endorsed by genetic [*e*.*g*., Cooper-Knock *et al*., 2014; Gómez-Tortosa *et al*., 2017], neuropathological [*e*.*g*., Geser *et al*., 2011; Kosaka *et al*., 2012] and neuroanatomofunctional [*e*.*g*., Tartaglia *et al*., 2009; Prudlo *et al*., 2012; De Vries *et al*., 2017] evidence.

Despite the severity of language impairment being moderately homogeneous across different MND phenotypes, it has to be noted that language deficits appeared to be more prevalent and severe in UMND patients when compared to LMND patients. This finding might be due to a greater cortical involvement in the former when compared to the latter group.

Mild-to-moderate lexical-semantic deficits proved to be highly prevalent in both UMND and LMND patients - in line with contributions investigating this component in classical ALS patients [*e*.*g*., Leslie *et al*., 2015]. Furthermore, mild morpho-syntactic deficits previously described in classical ALS patients [*e*.*g*., Kamminga *et al*., 2016] can be similarly detected in patients diagnosed with UMND/LMND phenotypes. Although rarely assessed in both classical ALS and other MND phenotypes, mild phonological deficits have been seldom detected in UMND patients – consistently with related contributions in classical ALS patients [*e*.*g*., Tstermentseli *et al*,, 2015].

Findings from studies in which cognitive screeners were administered appeared to be as well consistent with those in which component-specific tests were adopted: both the ACE-R/-III and the ECAS linguistic sub-tests proved to be able to detect language dysfunctions in both UMND and LMND patients.

With respect to lexical-semantic components, it is worthy to note that a systematic object-action/noun-verb difference in naming abilities was systematically detected in both UMND and LMND patients. This finding is consistent with evidence regarding classical ALS patients – who also are acknowledged to find it more difficult to name actions/verbs rather than objects/nouns [*e*.*g*., Papeo *et al*., 2014; York *et al*., 2014]. It can be thus speculated that, consistently with current guidelines for cognitive assessment in ALS patients [Strong *et al*., 2017; Woolley & Rush, 2017], action-naming tasks might represent a sensitive tool for detecting language changes in UMND/LMND patients too.

As far as written language is concerned, central dysgraphic features - likely due to an involvement of the graphemic buffer [Zago *et al*., 2008] - have been reported in patients with UMND, whereas not in those with LMND. With this regard, it is worth noting that writing errors found in French and Italian PLS patients is partially overlapping with those reported by Ferguson & Boller (1977) in English ALS patients – *i*.*e*., spelling and grammatical errors. Moreover, it has to be noted that no evidence of writing disorders in LMND patients have been provided.

It is worth noting that several case reports/series have documented that UMND patients can present with PPA [Östberg & Bogdanovic, 2011; Gazzulla *et al*., 2019] – similarly to what has been reported in ALS patients [Tan *et al*., 2019]. It can be thus reasonably inferred that language deficits might dispose along a *continuum* in MND phenotypes different from classical ALS too [Strong *et al*., 2017; Pinto-Grau *et al*., 2018].

### 4.2. Methodological considerations

First, it has to be noted that patients that are initially diagnosed with a MND phenotype different from classical ALS can be diagnosed with classical ALS over time [Cortés-Vicente *et al*., 2017]. Therefore, due to the challenges of MND phenotyping [Al-Chalabi *et al*., 2016], it cannot be ruled out that the underlying pathology of MND patients that have been classified in this work as different-from-classical-ALS was the same of classical ALS itself [Turner, 2019]. Furthermore, it must be taken into consideration that, in some of the studies included [*e*.*g*., Gallassi *et al*., 1985; Consonni *et al*., 2013; Spinelli *et al*., 2016; 2019], a considerable within-cohort heterogeneity could be detected – *e*.*g*., patients falling under the same classification (*e*.*g*., LMND) but affected with different MND phenotypes (*e*.*g*., prLMND, PMA, flail arm syndrome, flail leg syndrome, respiratory ALS) being included in the same cohort [Spinelli *et al*., 2016]. The aforementioned considerations suggest that caution should be exerted when generalizing results regarding a specific MND phenotype.

With regard to disease-related outcomes, it is of interest to note that dysarthric speech has been only seldom reported in the considered patients [*e*.*g*., Caselli *et al*., 1995; Zago *et al*., 2008] and also happened to be an exclusion criteria for patients to be recruited [Gallassi *et al*., 1985]. This suggests that reports of language impairment in this work should not have been distorted by dysarthria [Cobble, 1998]. Furthermore, FTD has been rarely reported among patients in the included studies [De Vries 2019a; 2019b] – this suggesting that language impairment in the vast majority of cohorts was not likely to be carried by the presence of severely-impaired/PPA patients.

It has also to be acknowledged that highly variable sample sizes have been detected in the included studies - ranging from small-to-medium samples [*e*.*g*., Zago *et al*., 2008; Raaphorst *et al*., 2015] to large cohorts [*e*.*g*., De Vries *et al*., 2019a]. Furthermore, it is worth noting that only one of the studies [Finegan *et al*., 2019] took into consideration a population-based cohort. With this regard, it must be also pointed out that some contributions did not assess language functioning in all the patients that were initially recruited [*e*.*g*., Caselli *et al*., 1995].

Moreover, not all studies included a control group. Despite comparisons with normative data being frequently provided, the absence of a control group might have led to an under-estimation of sub-clinical/mild language deficits [Pinto-Grau *et al*., 2018]. Indeed, the presence of severe language impairment/a full-blown PPA in MND patients has to be regarded as infrequent when compared to mild-to-moderate language deficits [Taylor *et al*., 2013; Pinto-Grau *et al*., 2018]. This last assertion is also supported by that fact that borderline deficits have been occasionally detected in the included studies [*e*.*g*., Raaphorst *et al*., 2011; 2015].

Moreover, it has to be considered that inferences regarding the neuro-anatomical/-functional correlates of language functions in patients with different-from-classical-ALS MND phenotypes can be hardly drawn based on the results of the present review – since only one study provides with specific correlations between language and neural measures [Canu *et al*., 2013]. Future studies might thus focus on assessing the association between language and neural measures in different-from-classical-ALS patients.

As far as assessment methods are concerned, it has to be acknowledged that language was predominantly assessed in the oral and productive modalities; similarly, at a component level, lexical-semantic abilities were the most frequently investigated. This might possibly lead to an over-estimation of deficits regarding these modalities/components. In the studies where either the ECAS or the ACE-R/-III were administered, analyses and generalizations of results with respect to the different linguistic components of interest have to be done with caution. Indeed, it must be noted that the score of the linguistic sub-test provide with a measure of global language functioning – *i*.*e*., do not provide with detailed information regarding which component/s might be affected. However, it is reasonable to infer that an abnormal performance on the aforementioned global suggests the presence of a language dysfunction.

Furthermore, it is not trivial to draw valid cross-linguistic inferences from a body of evidence characterized with high heterogeneity as far as the spoken language is concerned [Pinto-Grau *et al*., 2018]. Indeed, different components/functions might be differently involved in different languages when language disorders are concerned [Canu *et al*., 2020].

Finally, it has to be taken into consideration that the actual presence of language deficits may be biased due to the lack of corrections for motor impairment [Abrahams *et al*., 2014; Pinto-Grau *et al*., 2018], which have not always been implemented - except for studies in which the ECAS was administered. This latter assertion points out the need for development of component-/function-level language tests with correction for motor disability (*i*.*e*., dysarthria and/or upper-limb impairment) [Abrahams *et al*., 2014; Pinto-Grau *et al*., 2018].

## 5. Conclusions

Overall, findings of this review suggest that language deficits can occur also in patients affected with different-from-classical-ALS MND phenotypes (UMND/LMND). The language profile of UMND/LMND patients appears to partially overlaps with that of classical ALS patients. Furthermore, patients affected with UMND phenotypes present with more severe language dysfunctions when compared to LMND patients. Mild-to-moderate lexical-semantic deficits appeared to be highly prevalent in both UMND and LMND patients, with a selective difficulty in action-vs. object-naming being systematically detected. This latter finding suggests that, consistently with guidelines for cognitive assessment in ALS patients, action-naming tasks might represent a valid and sensitive tool for assessing language in UMND/LMND patients too. By contrast, morpho-syntactic and phonological components happened to be found as mildly impaired - mostly in UMND patients. Central writing deficits have also been reported in UMND patients. It is thus reasonable to hypothesize that, as far as language dysfunction is concerned, patients with different-from-classical-ALS MND phenotypes could be validly included the MND-FTD *continuum* at a neuropsychological level.

## Data Availability

No datasets are associated with this manuscript.

## Disclosure statement

The Authors have no known conflicts of interest to declare.

## Fundings

This work did not receive any specific funding.

## Notes

### Competing Interest Statement

The authors have declared no competing interest.

### Funding Statement

This study did not receive any specific funding.

### Author Declarations

No Ethical approval was needed for this study to be implemented since it is a systematic review on already published data.

